# Do recent family physician graduates practice differently? A longitudinal study of primary care visits and continuity in four Canadian provinces

**DOI:** 10.1101/2022.03.11.22272161

**Authors:** David Rudoler, Sandra Peterson, David Stock, Carole Taylor, Drew Wilton, Doug Blackie, Fred Burge, Rick Glazier, Laurie Goldsmith, Agnes Grudniewicz, Lindsay Hedden, Margaret Jamieson, Alan Katz, Adrian MacKenzie, Emily Marshall, Rita McCracken, Kim McGrail, Ian Scott, Sabrina Wong, M. Ruth Lavergne

## Abstract

**Background:** Complaints about lack of access to family physicians (FPs) has led to concerns about the role recent physician graduates have had in changes in the supply of primary care services in Canada. This study investigates the impact of career stage, time period, and graduation cohort on family physician practice volume and continuity over two decades.

**Methods:** Retrospective-cohort study of family physician practice from 1997/98 to 2017/18. Administrative health and physician claims data were collected in British Columbia, Manitoba, Ontario, and Nova Scotia. The study included all physicians registered with their respective provincial regulatory colleges with a medical specialty of family practice and/or billed the provincial health insurance system for patient care as family physicians. Median polish analysis of patient contacts and physician-level continuity was completed to isolate years-in-practice, period, and cohort effects.

**Results:** Median patient contacts per provider fell over time in the four provinces examined. In all four provinces, median contacts increased with years in practice until mid-to-late-career and declined into end-of-career. We found no relationship between graduation cohort and practice volume or FP-level continuity.

**Interpretation:** Recent cohorts of family physicians practice similarly to predecessors in terms of practice volumes and continuity of care. Since FPs of all career stages show declining patient contacts, system-wide solutions to recent challenges in the accessibility of primary care in Canada are needed.

---

While about 85% of Canadians have access to a regular primary care provider, timely access to primary care services in Canada falls short of comparable countries.^1^ Despite a growing supply of family physicians (FPs),^2^ there is concern that FPs practice has changed over time, which has had adverse effects on access to care. For instance, there is evidence that the volume of patient contacts and practice sizes are in decline^3,4^ and that FPs are less likely to provide comprehensive care compared to previous years.^5^

It has been asserted that the preferences and motivations of the current generation of early-career FPs are contributing to changes in family practice in Canada. For instance, some have argued that recent cohorts think more about work-life balance, are less career motivated, desire more mentorship, and are less likely to engage in comprehensive family practice.^6–10^

This paper measures trends in FP practice volume and continuity of care over two decades. A common method for investigating changes in population-level trends is age-period-cohort modeling. This approach involves understanding the separate impacts on service provision of an individual’s life cycle (age effects); shifts in sociodemographic, economic, and political contexts (period effects); and different choices made by groups entering practice at different times (cohort effects).

Understanding the relative magnitude of these age, period, and cohort effects is important for informing policy responses. Age effects imply that workforce planning should account for the practice decisions that FPs tend to make at different stages of their careers. Period effects would suggest the need for policy responses to sociodemographic, economic, and political events shaping the human resource market. Cohort effects would suggest the need for intervention during the formative stages of physician training.

## Methods

### Setting

This study uses administrative health data for four provinces: British Columbia, Manitoba, Ontario, Nova Scotia. Since healthcare services fall under provincial and territorial jurisdiction, each province and territory can establish its own policies with respect to health human resource planning, payment, and practice models. FPs are compensated by provincial health insurance systems. The majority of Canadian FPs practice privately and are paid fee-for-service, but the proportion of FPs in this model delivery and payment varies across the country.^11–13^

### Data

We used linked administrative health databases housed in BC (PopDataBC), Ontario (ICES), Manitoba (Manitoba Centre for Health Policy), and Nova Scotia (Health Data Nova Scotia). We accessed comparable databases, developed comparable definitions for all variables and conducted parallel analyses. Databases accessed for this study included: registry files from provincial regulatory colleges, physician billing information, and patient registration file for provincial insurers. Note that billing data includes fee-for-service billing and shadow billing information in each province.^14–17^ We are required to note that Ontario datasets were linked using encoded identifiers and analyzed at ICES. Ethics approval has been obtained from the UBC-SFU Harmonized Behavioural Research Ethics Board (Ethics #: H18-03291), University of Ottawa Ethics Board (Ethics #: S-05-18-776), Ontario Tech University Ethics Board (Ethics #: 14867), Nova Scotia Health Authority Ethics Board (Ethics #: 1023561), and the University of Manitoba (Ethics # HS23897 (H2020:208)). Further details on these data were published as part of the study protocol.^18^

We included all physicians registered with their respective provincial regulatory colleges with a medical specialty of family practice and/or billed the provincial health insurance system for patient care between the 1997/98 and 2017/18 fiscal years. We excluded physicians in any fiscal year where they had fewer than 100 unique patient-day contacts or had fewer than 50 days during the year where they billed for services delivered. We also excluded physicians in any fiscal year where they had a specialty other than family practice.

### Age, Period and Cohort

Rather than biological age, we used years in practice as our measure of ‘age, which was defined as fiscal year minus graduation year. We subtracted two years from years in practice to account for time in residency and any billings made during those two years were excluded. Year of graduation was not available in Manitoba, so the analysis relied on the year an FP first registered with the provincial insurer. ears in practice in that province were truncated at 23 years as the first observed year of registration was 1973. Period was defined as the current fiscal year (from 1997/98 to 2017/18). Cohort was defined as the current fiscal year minus years in practice.

### Outcomes

Outcomes were median annual patient contacts and median annual physician-level continuity as reported by physicians through their billing records. The medians were used as the distributions of physician-level patient contacts and continuity were right-skewed. Patient contacts represented unique patient-physician-date combinations in physician billings for service delivered in-person, or virtually. Contacts excluded laboratory services, imaging services, and no-charge referrals. Physician-level continuity was defined as the proportion of total annual contacts (excluding ED visits) that all patients seen by an FP had with that FP. For example, if over a fiscal year an FP saw two patients two times each and each patient had five FP visits in total, the continuity measure would equal (2+2)/(5+5) = 0.4.

### Other Variables

We also tracked physician sex and practice location (urban/rural). Sex is a binary variable self-reported by physicians at time of registration, and whether legal sex, sex assigned at birth, or gender is being reported cannot be confirmed. We also tracked the location practice (which could change over time), of medical degrees (Canada, international or unknown), billing days per year, contacts per billing day, unique patients seen, and the number of physicians with one or more shadow billings and one or more contacts in ambulatory locations. Shadow billings included instances where a FP billed a code for tracking purposes but did not receive full fee-for-service payment (see supplementary materials for further details).

### Statistical Analysis

Keyes et al. argue that age-period-cohort modeling requires a core assumption of whether a cohort is defined as a first-order effect that represents the unique conditions that shape life-long preferences (e.g., the cohort of people born soon after WWII experiencing a common set of experiences over their life course), or as the interaction between period and age (e.g., emerging theories of effects of the COVID-19 pandemic on school-age children specifically).^19^ This choice is conceptual, not empirical.^19^ We adopted the latter definition, contending that our study cohorts are best defined by non-equivalent period effects on FPs at different career stages. That is, the larger social and political context (“period”) is likely to affect different age groups differently.

We used the median polish approach^20^ that estimates second-order cohort effects.^19^ This approach uses a contingency table with the number of rows equal to the number of years in practice categories and the number of columns equal to the number of periods. We used equal three-year categories of period and years in practice. Thus, each cell of the contingency table contained the observed outcome for the corresponding years in practice-period combination (e.g., years in practice =3 to 5 years, period = FY2000 to FY2002). We regressed the outcomes on indicators for years in practice and period. Then, we used median polish, which iteratively subtracted row and column medians from the cell values, until the row and column medians approached zero. The residuals that remained in the cells were then regressed on cohort indicators. The regression coefficients were estimated using a linear model, and bootstrap standard errors with 1,000 iterations were generated to calculate 95% confidence intervals. The coefficient estimates and confidence intervals were plotted. Since all independent variables were categorical, a reference category was chosen for period (FY1997 to FY1999), years in practice (0 - 2 years), and cohort (FY1991 to FY 1993). The plots show a horizontal solid line at zero, which indicates no difference from the reference category. The plots for the main effects are provided in the following section, while stratified analyses for sex and rural/urban practice are provided in the supplementary materials.

## Results

Table 1 shows descriptive statistics for included FPs in all provinces at the beginning and end of the study period: fiscal years 1997/98 and 2017/18. Mean contacts billed by FPs declined in each province between the start and end of the study period, as did reported contacts per billing day and unique patients reported by the physician as being seen. The proportion of FPs using shadow billing increased in all provinces concurrently with the increase in the proportion of FPs compensated via alternative payment models.^13^ Physician-level continuity as indicated by physicians’ billing records remained stable.

**Table 1.**
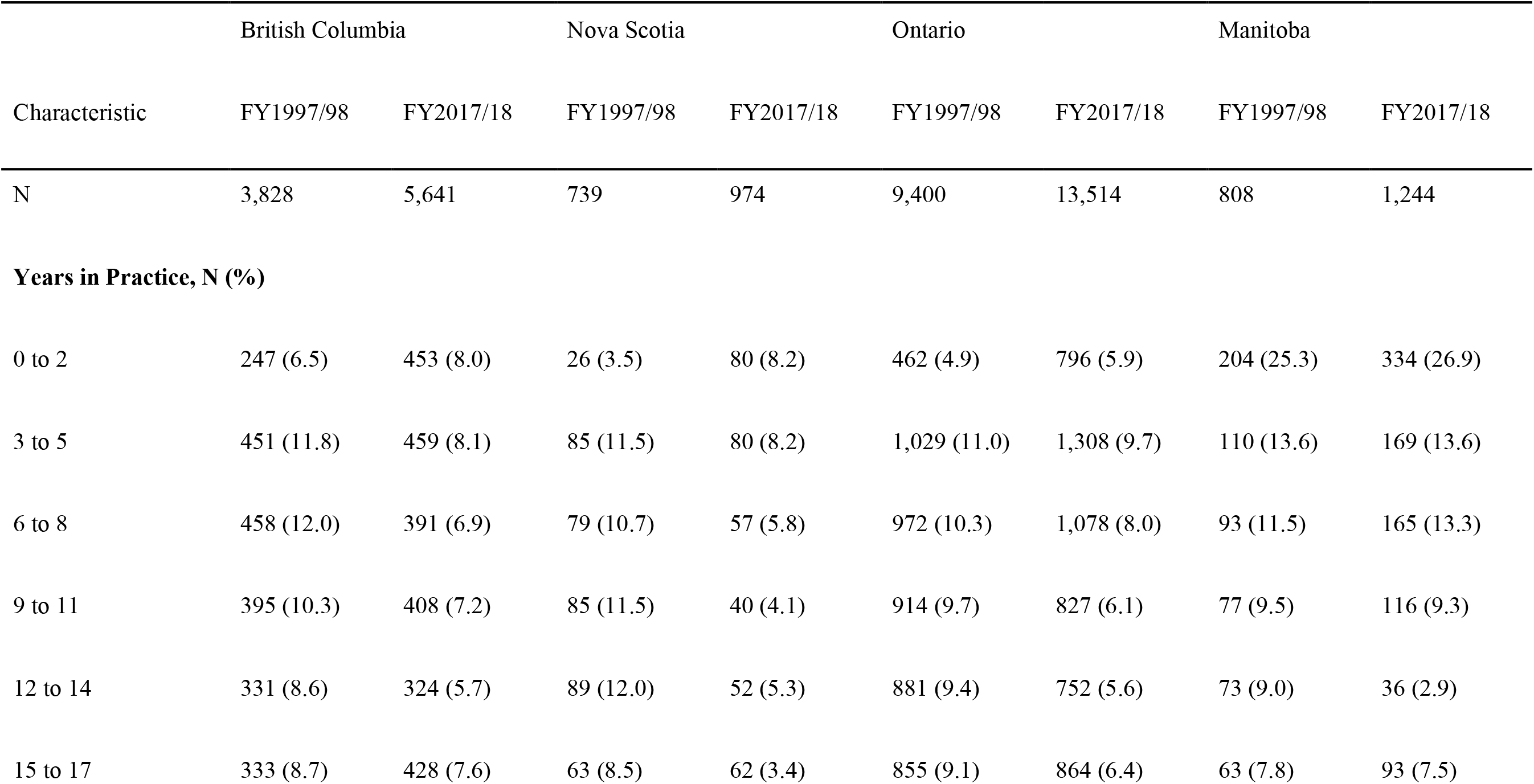
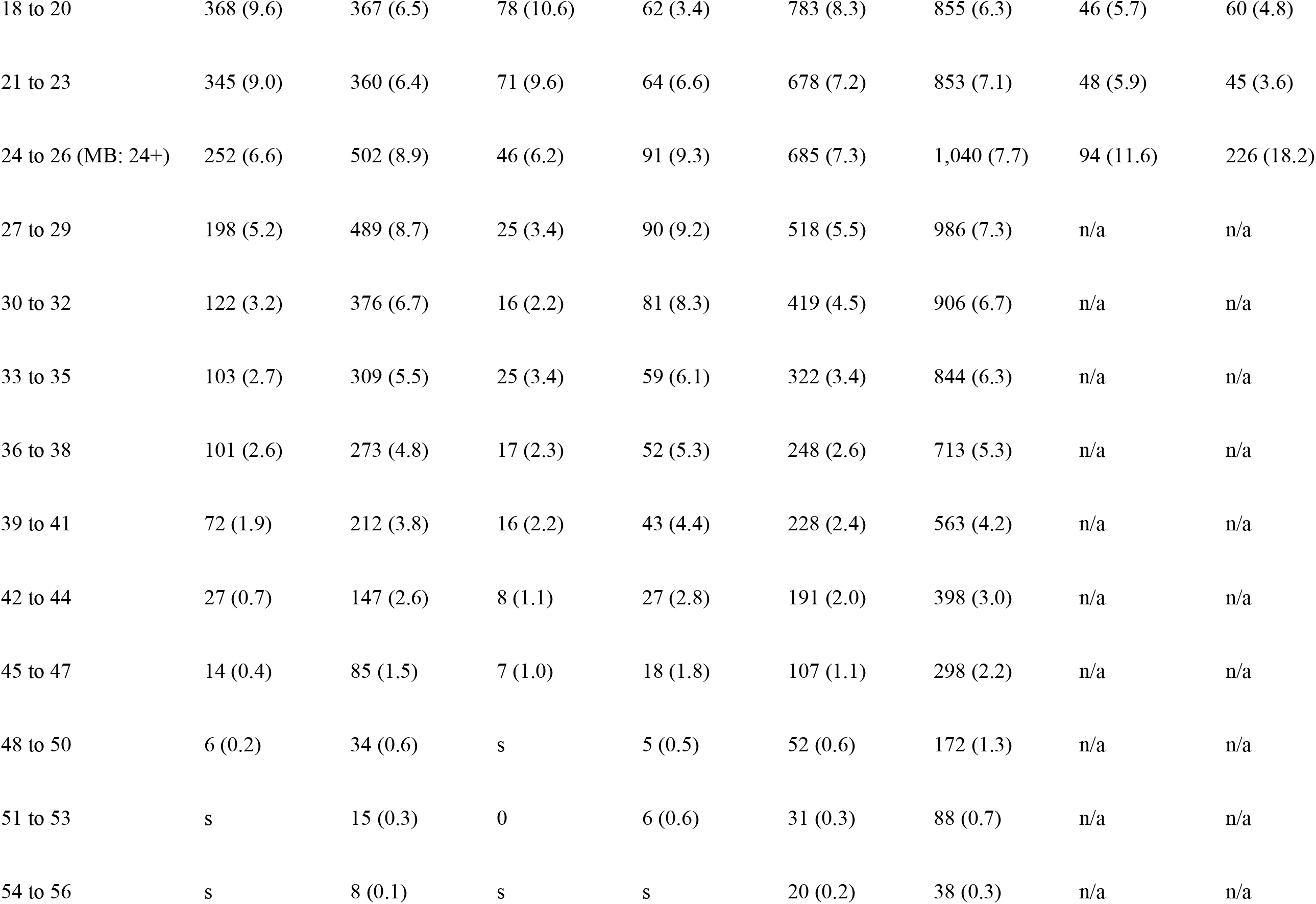
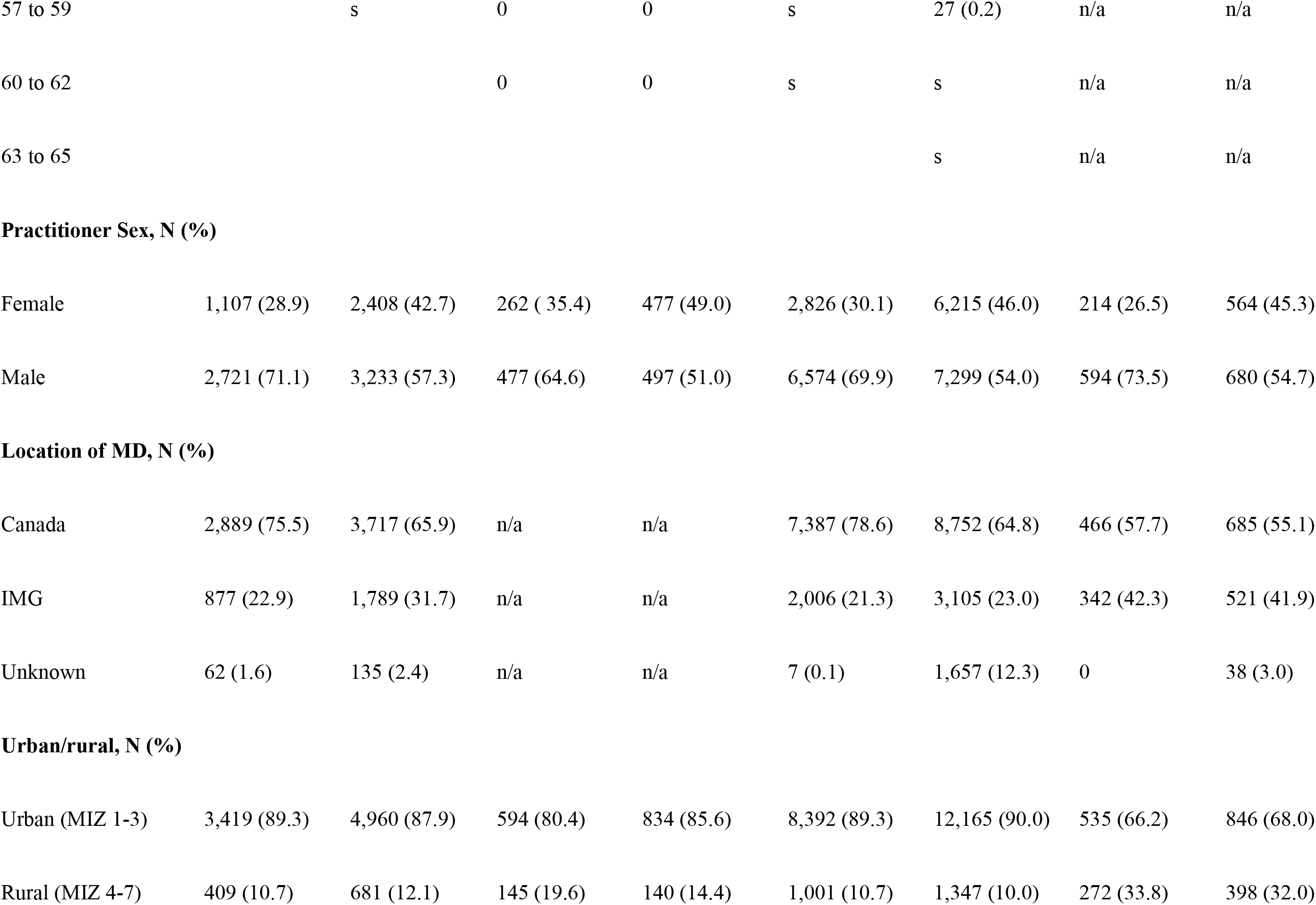
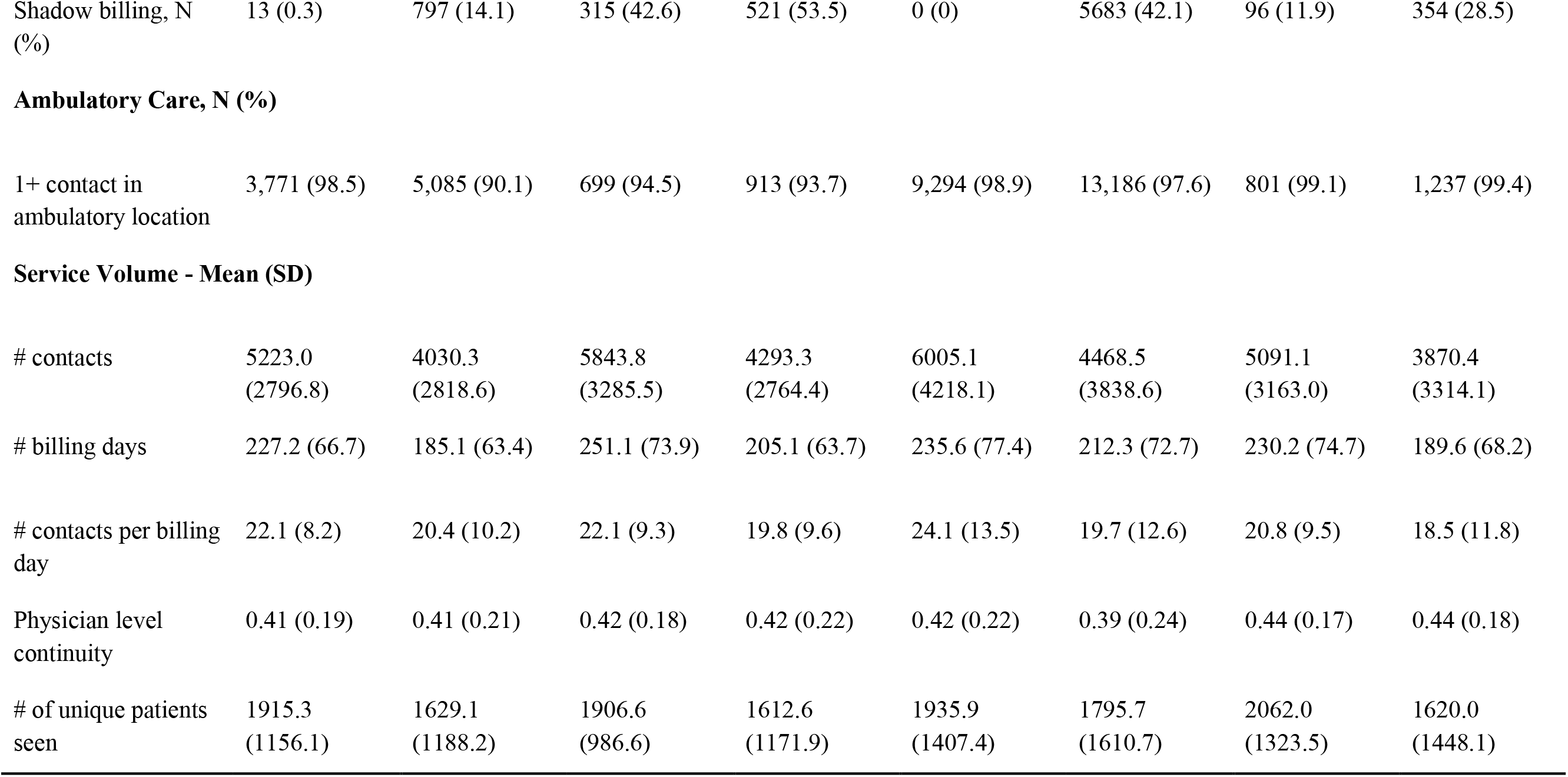
Physician characteristics, 1997/98 and 2017/18.

### Period

Figure 1 shows the effects of period on patient contacts. Relative to the earliest period (FY1997 to FY1999), median billed contacts declined over the study period. The pattern of decline is visible in all provinces, and the decline was more pronounced in rural practices and among male physicians.

**FIGURE 1.**
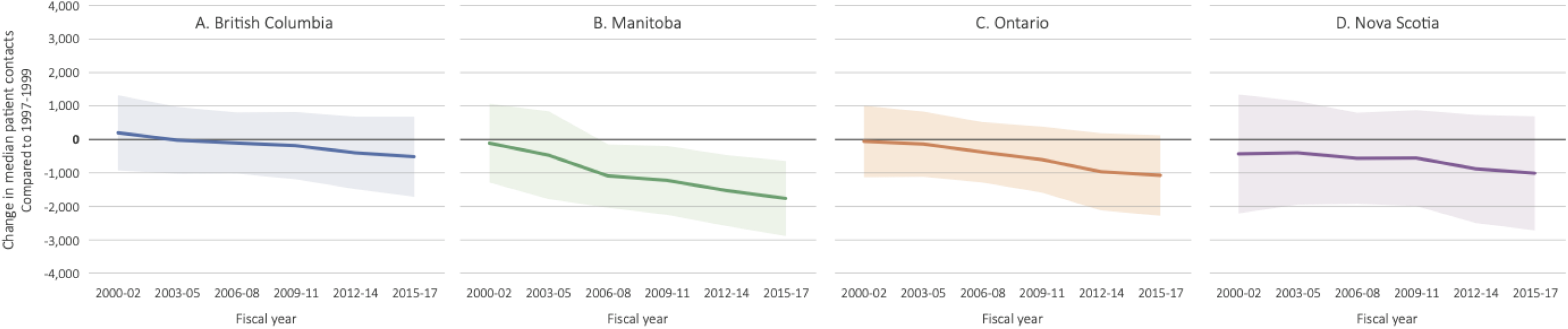
Period effects on median patient contacts, with 95% confidence intervals

Figure 2 shows the effects of period on median physician-level continuity, which remains stable over the study period. We do not observe clear differences when the data are stratified by FP sex or into rural and urban practice.

**FIGURE 2.**
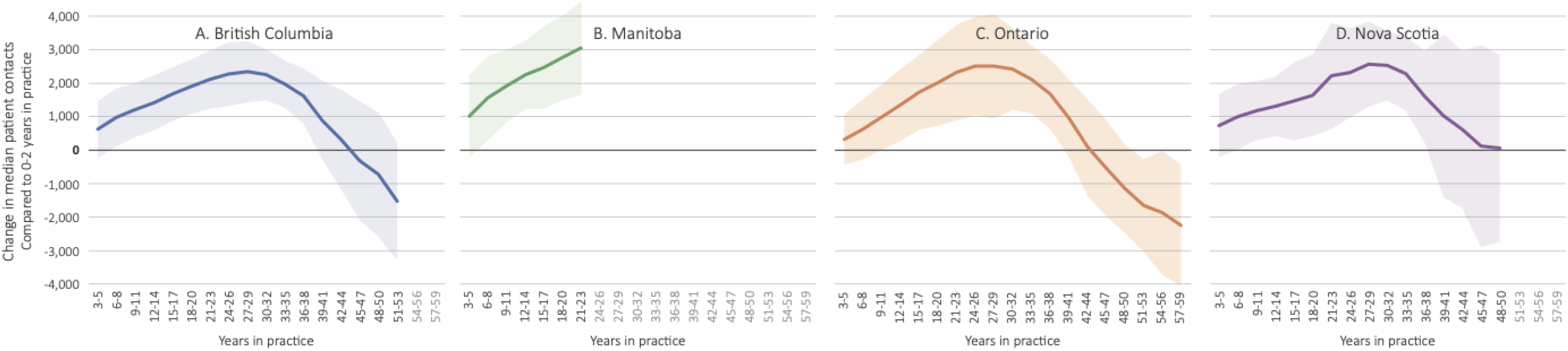
Years in practive effects on median patients contacts, with 95% confidence intervals

### Years in practice

Figure 3 shows the effects of years in practice on median patient contacts in the four provinces. In all provinces, we clearly see the inverted-U-shaped curve indicating patient contacts increase year to year until 20+ years in practice before beginning to decline (point estimates are relative to 0-2 years in practice). Even in Manitoba where the data were truncated, we can see the beginnings of that same trend emerge. In the other provinces, median contacts peak at around 27 to 29 years, and then start to decline into end-of-career. The peaks of this trend are slightly higher for male physicians.

**FIGURE 3.**
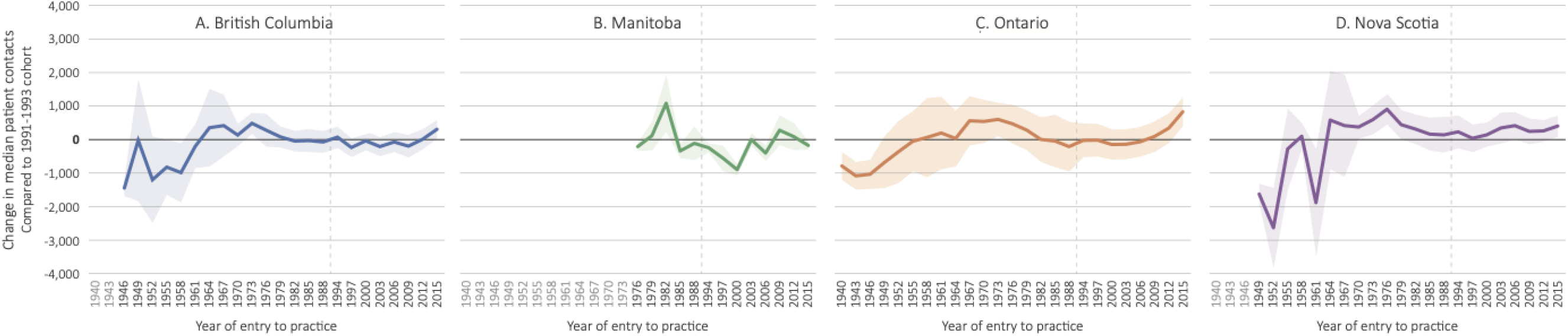
Cohort effects on median patients contacts, with 95% confidence intervals

Figure 4 shows the effects of years in practice on physician-level continuity. Again, we observe an inverted-U-shaped curve, but do not see declines in late-career in Ontario and Nova Scotia; instead, continuity in those provinces remains stable from mid to late-career.

**FIGURE 4.**
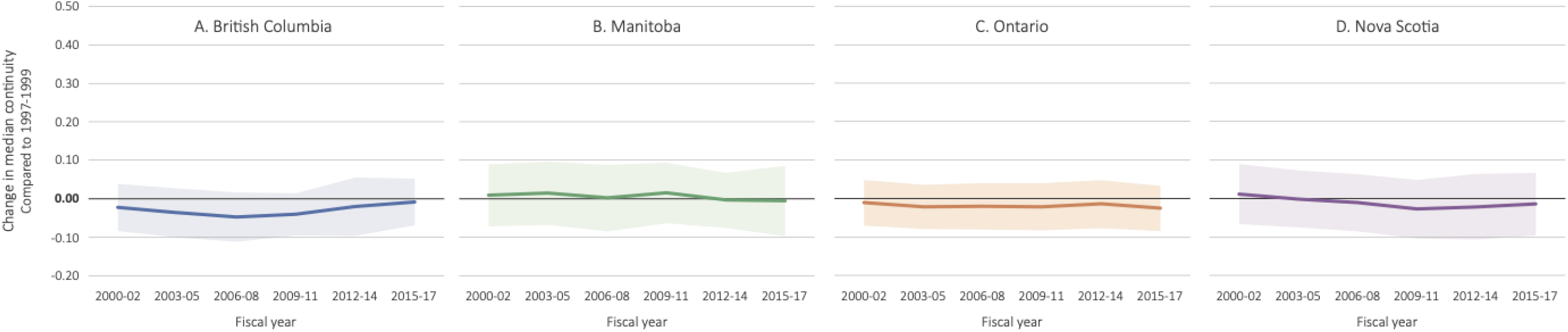
Period effects on median physician-level continuity, with 95% confidence intervals

### Cohort

Figure 5 shows the effects of cohort on patient contacts in the four provinces. Generally, the figures show no difference in billed patient contacts per year across cohorts, with the exceptions of the extremes of the cohort distribution. In three of the four provinces, FPs who started practice in the late 1940s and early 1950s (Manitoba data was not available for these cohorts) had lower median patient contacts than FPs who started practice in 1991 to 1993. Meanwhile, FPs who started practice in the mid to late 2010s had higher median patient contacts than FPs who started practice in 1991 to 1993.

**FIGURE 5.**
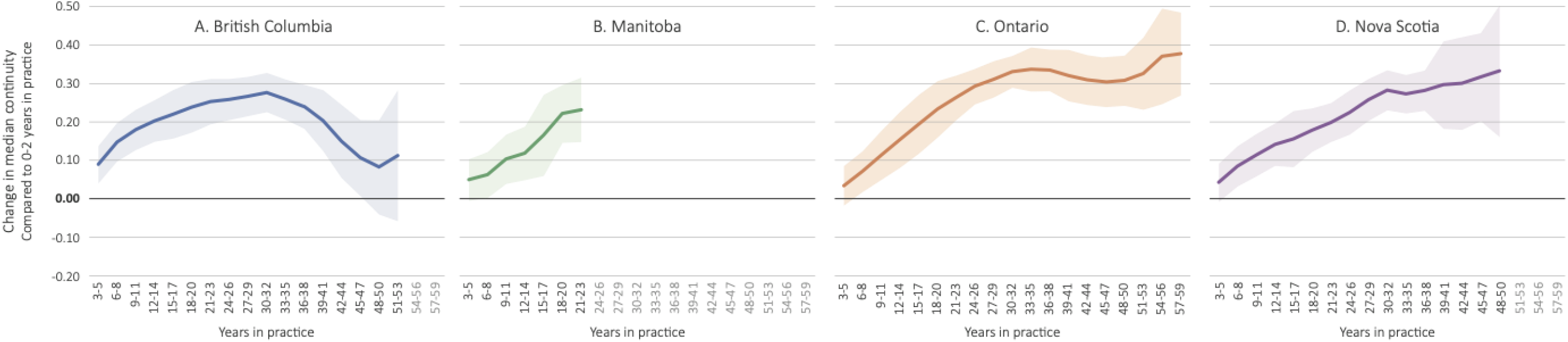
Years in practice effects on median physician-level continuity, with 95% confidence intervals

Figure 6 shows the effects of cohort on physician-level continuity in the four provinces. The results are mostly consistent with those for patient contacts, with little difference in continuity by cohort in any province. Continuity was higher for FPs in Ontario in practice since the 1940s. Stratified analysis in that province revealed that there were a small number of senior urban FPs providing very high continuity of care.

**FIGURE 6.**
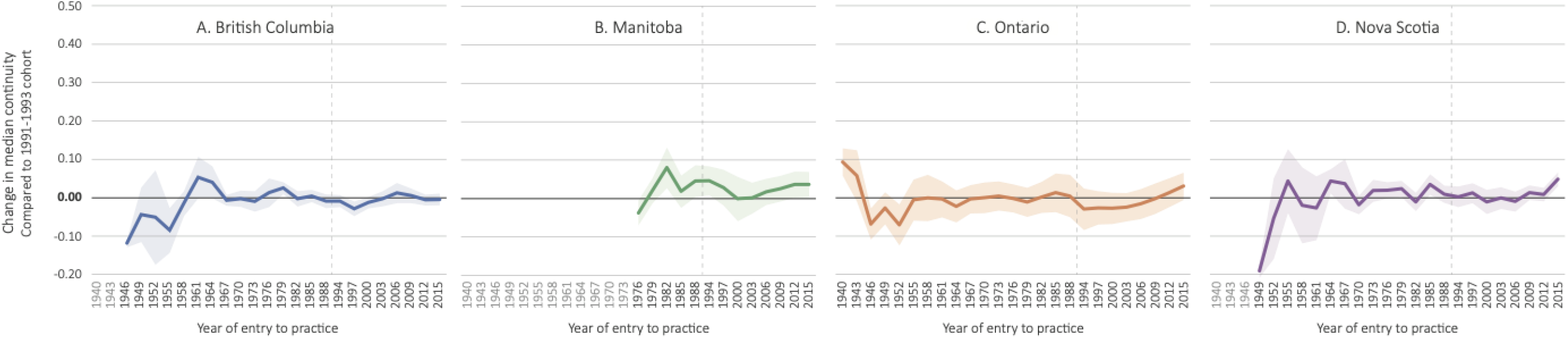
Cohort effects on physician-level continuity, with 95% confidence intervals

## Interpretation

In this longitudinal study of FP practice in Canada, we found that the median contacts fell over time in the four provinces examined. Meanwhile, in all four provinces median contacts increased with years in practice until mid-to-late-career and declined into end-of-career. These findings align with our prior expectations and with previous research.^3,4,21^ Similar to contacts, continuity increased with years in practice and fell in the later stages of a career in British Columbia, but not in Ontario or Nova Scotia. We found that patient contacts and continuity were not affected by the cohort that an FP was part of. Changes in FP practice over time were observed across physicians at all career stages, not just in those entering practice in recent years. Stated another way, while younger and older physicians bill for fewer patients than mid-career physicians, there is a general decline over each cohort of all physicians billing for fewer patients, and not a disproportionate reduction in recent graduates compared to past graduates.

In previous studies that used physician survey data, Watson et al. found that younger FPs had smaller workloads in 2003 than their peers did ten years earlier, perhaps suggesting the existence of a cohort effect.^4^ Using similar data but different methods, Crossley et al. and Sarma et al. found no evidence of cohort effects.^22,23^ Our replication of these findings is interesting given we used a different measure of supply — unique patient contacts rather than self-reported hours of patient care — and applied different empirical methods.^22,23^

We cannot say whether declines in patient contacts reflect an increase in patient complexity, an increase in administrative burden, increased quality, or different choices about work across all primary care physicians; we can, however, say that these observed declines are not unique to current early career physicians. While further work is needed to parse out the causal mechanisms for declines in service volume, our findings do suggest several important considerations for decision-makers. First, because practice patterns differ over the course of physicians’ careers, it is important to pay attention to the age distribution of the physician workforce in health workforce planning. For example, these results suggest that a physician workforce with more physicians at the extremes of the distribution will bill for lower quantities of service compared to a distribution with the majority of physicians in mid-career.^24^ Put another way, these results suggest that a workforce made up mostly early- and late-career physicians may be able to care for a smaller population of patients than a workforce made up of mostly mid-career physicians, other things being equal. Second, the decline in reported billings across all physician cohorts in each province indicates that, even with increasing per-capita supply of FPs, additional resources will be needed to maintain or improve access to primary care, other things being equal. Third, since FPs of all career stages have decreasing supply, solutions to recent declines in service quantity will need to be broadly targeted and system-wide, rather than focused on a specific cohort of FPs. Fourth, since continuity rises and falls similarly to quantity of service, then seeking interventions to increase continuity early in a FP career may support improved continuity of the practice life of a FP and these policies, if implemented, should be evaluated.

Our study had several limitations. First, this study used billing data that do not fully capture the scope or complexity of the services. Neither levels of service received by patients nor workload of family physicians can be measured comprehensively with billing data. Second, observed patient contacts may have been affected by increases in alternative payment plans (APPs) and shadow billing over the study period in all physician cohorts. These may, at least in part, account for observed declines in patient contacts but are unlikely to have “masked” any reductions in service volume in recent graduates. Further, the fact that there was such similarity in the observed effects across provinces despite substantial differences in APP uptake between them suggests that the influence of shifting to APPs on these relationships may be relatively small. Third, we defined cohort as an interaction between period and years in practice which facilitated robust estimation of this effect as a non-additive combination of these factors. Our results are consistent with previous studies that relied on the alternative definition of cohort,^23^ but future analyses of longitudinal administrative data of physician practice could determine if different definitions produce different results.

## Conclusion

This study analyzed generational effects on FP practice across four Canadian provinces. Declines in service volume as reported by physicians through their billings were observed in all provinces, with expected trajectories of service volume and continuity over a FPs’ career. We found no generational differences in FP practice. These findings are important for health workforce planning in primary care sectors across the country, and for the general discourse concerning the behaviours and preferences of recent medical graduates. Our findings highlight that intergenerational tension and blame is unfounded and only distracts from important issues in workforce planning in primary care sectors.

## Supporting information

STROBE Checklist

## Data Availability

The datasets from this study is are held securely in coded form at the data centres in each province. While legal data sharing agreements between these data centres and data providers (e.g., healthcare organizations and government) prohibit the data centres from making the dataset publicly available, access may be granted to those who meet pre-specified criteria for confidential access. For instance, for Ontario data these details concerning access are available at www.ices.on.ca/DAS. The full dataset creation plan and underlying analytic code are available from the authors upon request, understanding that the computer programs may rely upon coding templates or macros that are unique to the data centres where the analytic datasets were generated and are therefore either inaccessible or may require modification.

## Acknowledgements

British Columbia: Data for this project was accessed through Population Data BC, PopData Project #: 19-044. All inferences, opinions, and conclusions drawn in this article are those of the authors, and do not reflect the opinions or policies of the Data Steward(s). Ontario: This study was supported by ICES, which is funded by an annual grant from the Ontario Ministry of Health (MOH). The opinions, results and conclusions reported in this paper are those of the authors and are independent from the funding sources. No endorsement by ICES or the Ontario MOHLTC is intended or should be inferred. Parts of this material are based on data and information compiled and provided by MOH and the Canadian Institute for Health Information (CIHI). Nova Scotia: The data (or portions of the data) used in this report were made available by Health Data Nova Scotia of Dalhousie University (#2017-EGM-001). Although this research / health service assessment analysis is based on data obtained from the Nova Scotia Department of Health and Wellness, the observations and opinions expressed are those of the authors and do not represent those of either Health Data Nova Scotia or the Department of Health and Wellness. Manitoba: The authors acknowledge the Manitoba Centre for Health Policy for use of data contained in the Manitoba Population Research Data Repository under project #2020-026 (HIPC# 2020/2021-10). The results and conclusions are those of the authors and no official endorsement by the Manitoba Centre for Health Policy, Manitoba Health, or other data providers is intended or should be inferred. Data used in this study are from the Manitoba Population Research Data Repository housed at the Manitoba Centre for Health Policy, University of Manitoba and were derived from data provided by Manitoba Health.

This study also received funding from the Canadian Institutes of Health Research (CIHR-Project Grant 155965).

## References

1. Canadian Institute for Health Information (CIHI). How Canada Compares: Results From The Commonwealth Fund’s 2016 International Health Policy Survey of Adults in 11 Countries [Internet]. Ottawa, ON: CIHI; 2017. Available from: https://www.cihi.ca/en/health-system-performance/performance-reporting/international/wait-times-for-primary-and-specialist

2. Canadian Institute for Health Information. Supply, Distribution and Migration of Physicians in Canada, 2020 - Data Tables. Ottawa, ON: CIHI; 2021.

3. Hedden L, Barer ML, McGrail K, Law M, Bourgeault IL. In British Columbia, The Supply Of Primary Care Physicians Grew, But Their Rate Of Clinical Activity Declined. Health Affairs. 2017 Nov;36(11):1904–11.

4. Watson DE, Slade S, Buske L, Tepper J. Intergenerational Differences In Workloads Among Primary Care Physicians: A Ten-Year, Population-Based Study. Health Affairs. 2006 Nov 1;25(6):1620–8.

5. Chan BTB. The declining comprehensiveness of primary care. CMAJ. 2002 Feb 19;166(4):429–34.

6. Glauser W, Tepper J. Can family medicine meet the expectations of millennial doctors? [Internet]. 2016 [cited 2017 Jun 6]. Available from: http://www.healthydebate.ca/2016/05/topic/young-doctors-family-medicine

7. Freeman TR, Boisvert L, Wong E, Wetmore S, Maddocks H. Comprehensive practice: Normative definition across 3 generations of alumni from a single family practice program, 1985 to 2012. Can Fam Physician. 2018;64(10):750–9.

8. Ladouceur R. What has become of family physicians? Can Fam Physician. 2012 Dec;58(12):1322.

9. Rowland K. The Voice of the New Generation of Family Physicians. Ann Fam Med. 2014 Jan 1;12(1):6–7.

10. Buddeberg-Fischer B, Stamm M, Buddeberg C, Klaghofer R. The new generation of family physicians--career motivation, life goals and work-life balance. Swiss Med Wkly. 2008 May 31;138(21–22):305–12.

11. Rudoler D, Peckham A, Grudniewicz A, Marchildon G. Coordinating primary care services: A case of policy layering. Health Policy. 2019 Feb;123(2):215–21.

12. Marchildon GP, Hutchison B. Primary care in Ontario, Canada: New proposals after 15 years of reform. Health Policy. 2016 Jul;120(7):732–8.

13. Canadian Institute for Health Information. National Physician Database, 2017-2018 - Data Release [Internet]. CIHI. 2019 [cited 2020 Jul 22]. Available from: http://secure.cihi.ca/estore/productFamily.htm?pf=PFC4052&lang=en$media=0

14. British Columbia Ministry of Health [creator](2018): Medical Services Plan Practitioner File. V2. Population Data BC [publisher]. Data Extract. College of Physicians and Surgeons of BC (2019). http://www.popdata.bc.ca/data

15. British Columbia Ministry of Health [creator](2018): Medical Services Plan (MSP) Payment Information File. V2. Population Data BC [publisher]. Data Extract. MOH(2019). http://www.popdata.bc.ca/data

16. Canadian Institute for Health Information [creator](2019): Discharge Abstract Database (Hospital Separations). V2. Population Data BC [publisher]. Data Extract. MOH(2019). http://www.popdata.bc.ca/data

17. British Columbia Ministry of Health [creator](2019): Consolidation File (MSP Registration & Premium Billing). V2. Population Data BC [publisher]. Data Extract. MOH(2019). http://www.popdata.bc.ca/data

18. Lavergne MR, Goldsmith LJ, Grudniewicz A, Rudoler D, Marshall EG, Ahuja M, et al. Practice patterns among early-career primary care (ECPC) physicians and workforce planning implications: protocol for a mixed methods study. BMJ Open. 2019 Sep 1;9(9):e030477.

19. Keyes KM, Utz RL, Robinson W, Li G. What is a cohort effect? Comparison of three statistical methods for modeling cohort effects in obesity prevalence in the United States, 1971–2006. Soc Sci Med. 2010 Apr 1;70(7):1100–8.

20. Tukey JW. Exploratory data analysis. Reading, Mass: Addison-Wesley Pub. Co; 1977. 688 p. (Addison-Wesley series in behavioral science).

21. Simkin S, Dahrouge S, Bourgeault IL. End-of-career practice patterns of primary care physicians in Ontario. Canadian Family Physician. 2019 May 1;65(5):e221–30.

22. Crossley TF, Hurley J, Jeon S-HS. Physician labour supply in Canada: a cohort analysis. Health Econ. 2009;18(4):437–56.

23. Sarma S, Thind A, Chu MK. Do new cohorts of family physicians work less compared to their older predecessors? The evidence from Canada. Social Science and Medicine. 2011;72(12):2049–58.

24. Hedden L, Lavergne MR, McGrail KM, Law MR, Cheng L, Ahuja MA, et al. Patterns of physician retirement and pre-retirement activity: a population-based cohort study. CMAJ. 2017 Dec 11;189(49):E1517–23.

25. Statistics Canada. Statistical Area Classification (SAC). 2015.

26. Schultz SE, Glazier RH. Identification of physicians providing comprehensive primary care in Ontario: a retrospective analysis using linked administrative data. CMAJ Open. 2017;5(4):E856–63.

