## Supplementary Materials for "Do recent family physician graduates practice differently? A longitudinal study of primary care visits and continuity in four Canadian provinces"

### Description of Variables

Practice location was assigned based on the Statistics Canada metropolitan influence zone of residence for patients seen by a FP. ^25^ The label of *urban* was applied if the majority of contacts occurred in zones 1 to 3, and *rural* if the majority of contacts occurred in zones 4 to 7. We also tracked the location of medical degrees (Canada, international or unknown), billing days, contacts per billing day, and unique patients seen. We also tracked the number of physicians with one or more shadow billings and one or more contacts in ambulatory locations. Shadow billings included instances where a FP billed a code for tracking purposes but did not receive full fee-for-service payment. To encourage FPs to continue to submit shadow codes, some provincial governments offered financial incentives,^26^ compared billings to expected service volumes, and conducted audits.

### Stratified Analysis

Figures A1 to A6 present the results of our age-period-cohort analysis stratified by sex. Note that this stratification employs a binary indicator for male and female. This information is self-reported by physicians at time of registration, and whether legal sex, sex assigned at birth, or gender is being reported cannot be confirmed. Recognizing this lack of specificity in our measures we describe these variables as sex.

**Figure A1 - Period Effects on Median Patient Contacts**

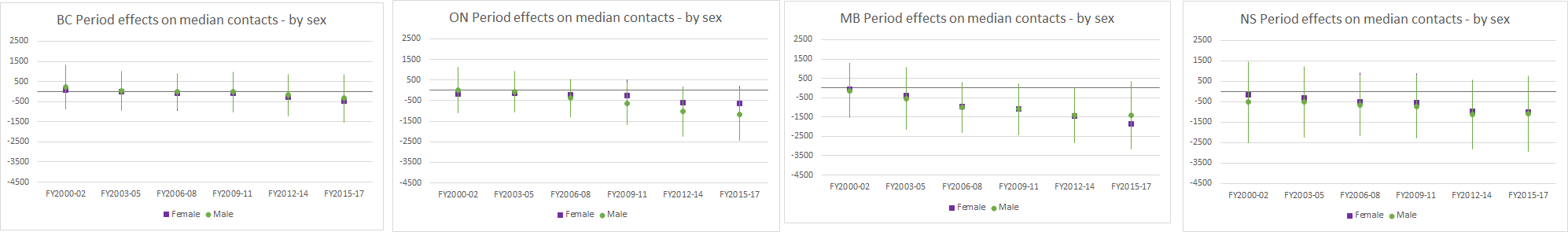

**Figure A2 - Years in Practice Effects on Median Patient Contacts**

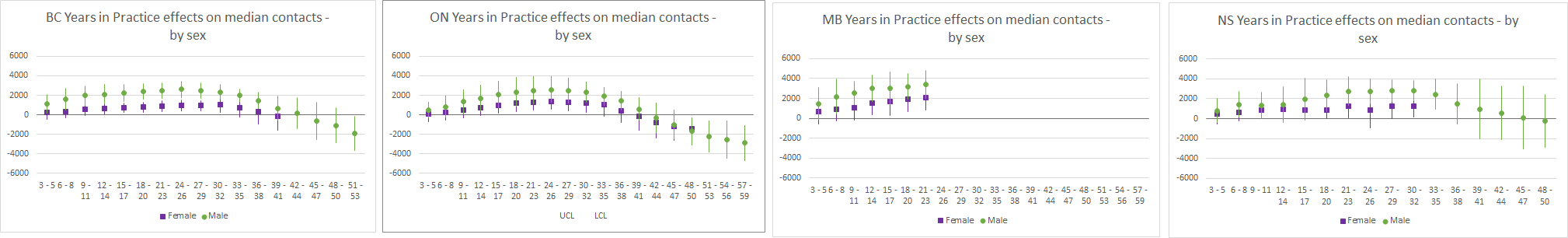

**Figure A3 - Cohort Effects on Median Patient Contacts**

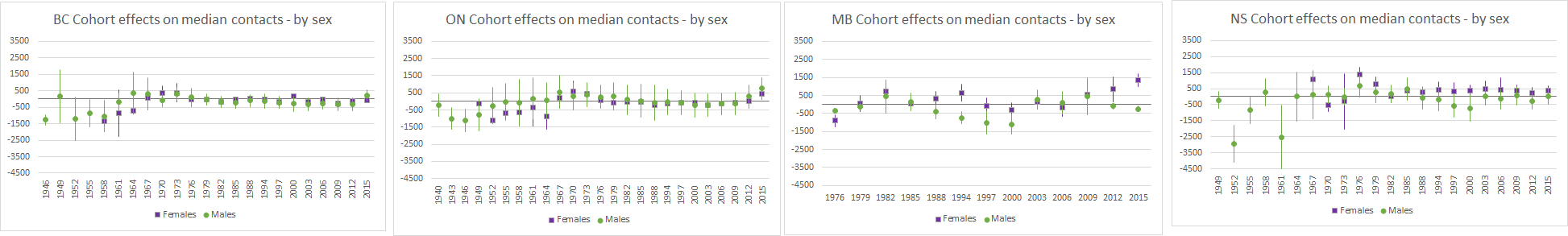

**Figure A4 - Period Effects on Median Physician-Level Continuity**

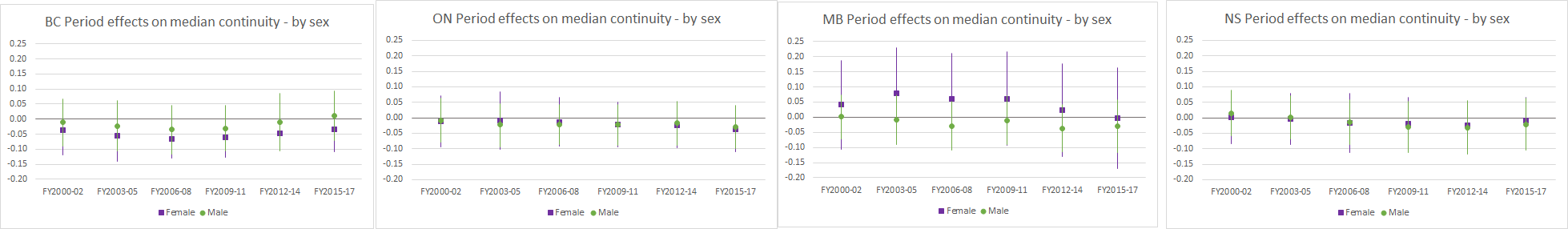

**Figure A5 - Years in Practice Effects on Median Physician-Level Continuity**

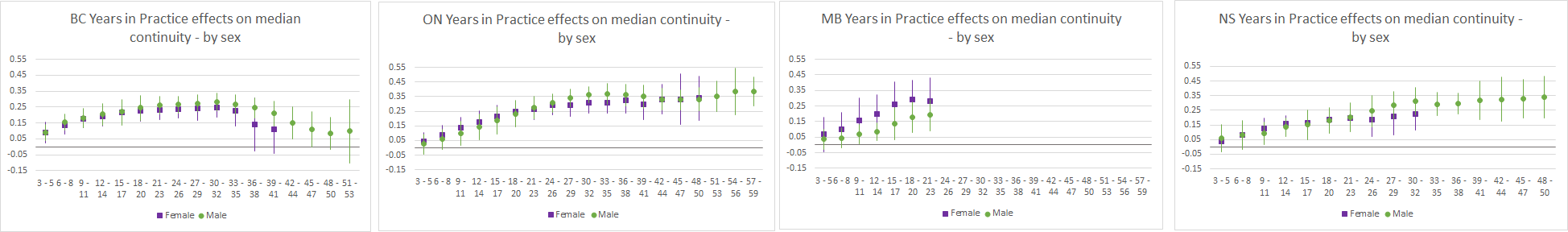

**Figure A6 - Cohort Effects on Median Physician-Level Continuity**

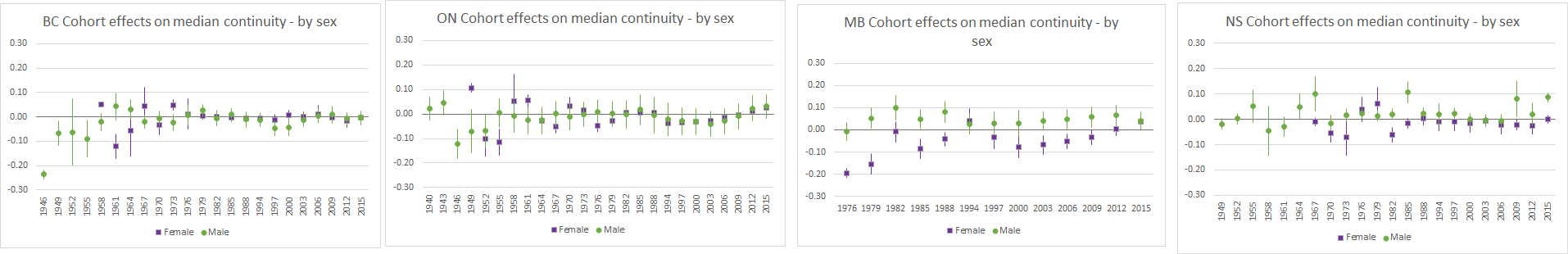

Figures A7 to A12 present the results of our age-period cohort analysis stratified by rural/urban.

**Figure A7 - Period Effects on Median Patient Contacts**

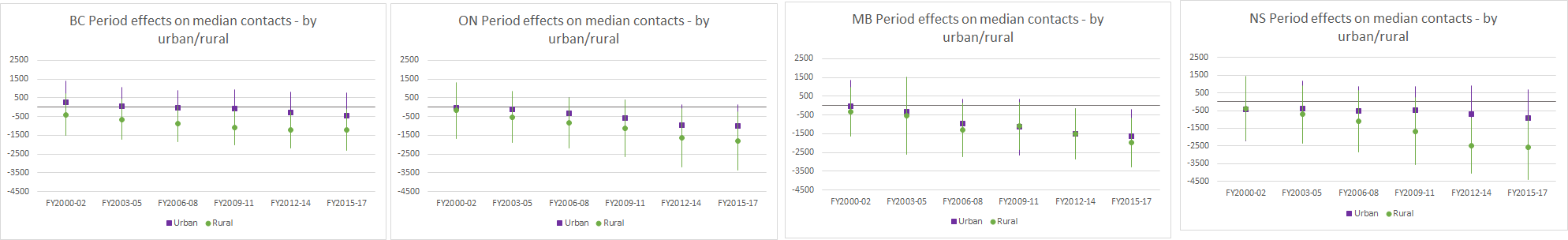

**Figure A8 - Years in Practice Effects on Median Patient Contacts**

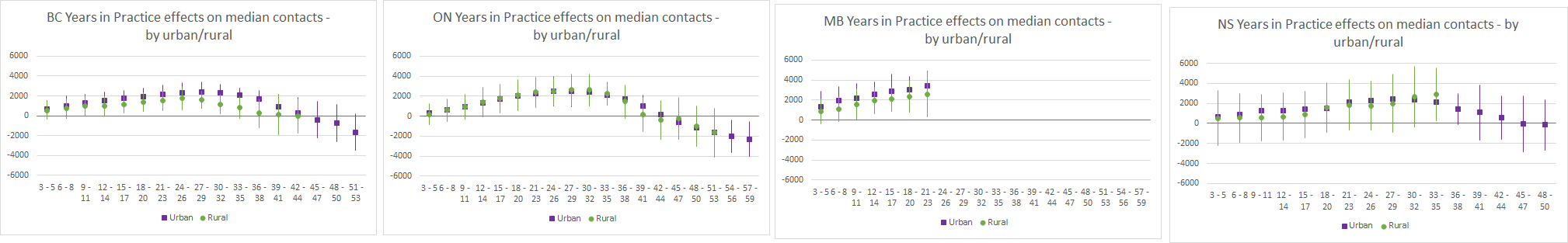

**Figure A9 - Cohort Effects on Median Patient Contacts**

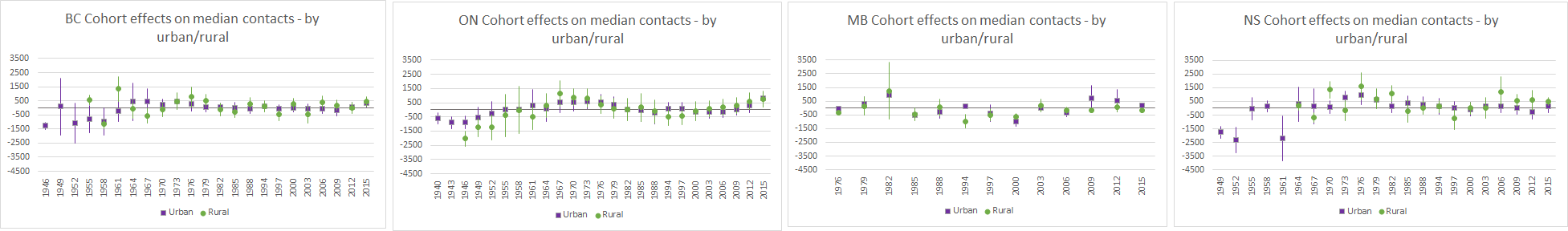

**Figure A10 - Period Effects on Median Physician-Level Continuity**

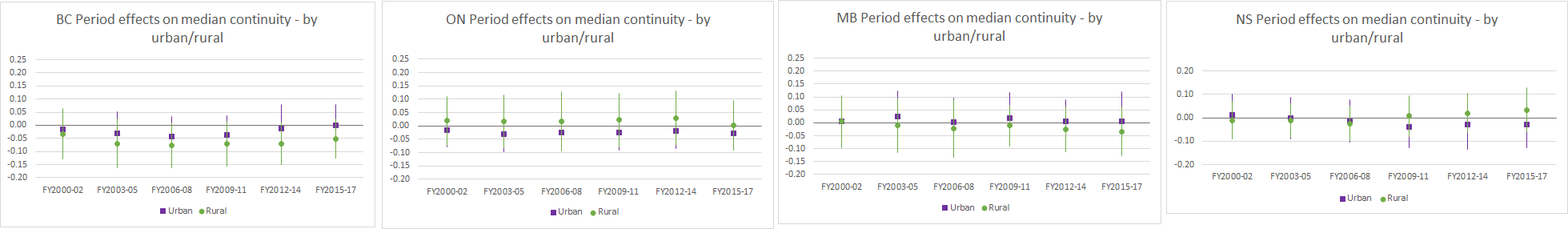

**Figure A11 - Years in Practice Effects on Median Physician-Level Continuity**

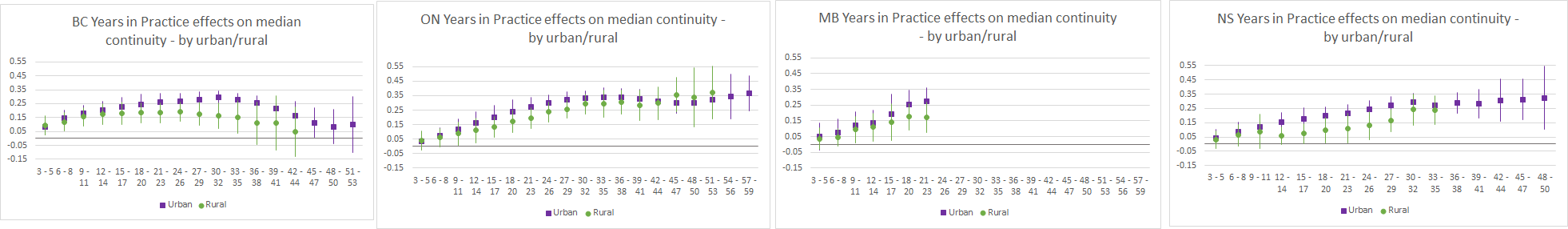

**Figure A12 - Cohort Effects on Median Physician-Level Continuity**

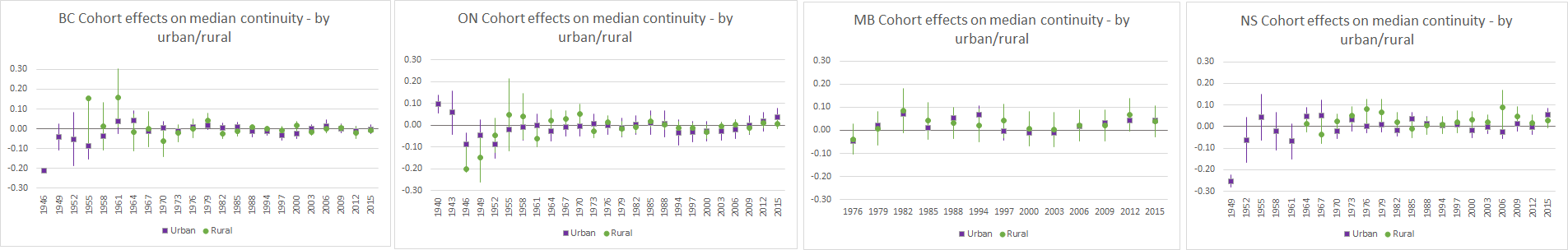

### Changes in Shadow Billing and Alternative Payment

Table A1 summarizes the changes in shadow billing by years in practice between fiscal years 1997/98 and 2017/18 and across all four study provinces.

| **Table A1 Shadow billing by years in practice** | | | |  |  |  |  |  |
| --- | --- | --- | --- | --- | --- | --- | --- | --- |
|  | British Columbia | | Nova Scotia | | Ontario | | Manitoba | |
| Shadow Billing | FY1997/98 | FY2017/18 | FY1997/98 | FY2017/18 | FY1997/98 | FY2017/18 | FY1997/98 | FY2017/18 |
| Years in Practice, N (%) |  |  |  |  |  |  |  |  |
| 0 to 4 | 7 (1.3) | 193 (25.1) | 50 (61.7) | 115 (83.3) | 0 | 527 (30.8) | 62 (23.7) | 194 (42.2) |
| 5 to 9 | s | 132 (19.7) | 69 (48.6) | 66 (72.5) | 0 | 797 (44.9) | 15 (8.9) | 85 (33.0) |
| 10 to 19 | s | 197 (16.2) | 107 (42.5) | 112 (60.9) | 0 | 1185 (43.8) | 11 (4.9) | 58 (24.3) |
| 20 to 29 (MB: 20+) | s | 168 (11.0) | 79 (45.9) | 123 (46.4) | 0 | 1410 (43.2) | 8 (5.3) | 17 (5.9) |
| 30 to 39 | 0 | 86 (8.2) | 6 (9.7) | 75 (37.1) | 0 | 1252 (46.8) | n/a | n/a |
| 40 to 49 | 0 | 20 (5.2) | s | 29 (35.4) | 0 | 464 (39.7) | n/a | n/a |
| 50+ | 0 | s | 0 | s | 0 | 48 (22.9) | n/a | n/a |
